# Processing negative autobiographical memories in a foreign language

**DOI:** 10.1101/2022.12.19.22283709

**Authors:** Isabel Ortigosa-Beltrán, Irene Jaén, Azucena García-Palacios

## Abstract

The use of a foreign language has been introduced in the clinical setting as a form of emotional distance to help deal with negative experiences. However, its evidence of reducing emotionality during the emotional processing is still scarce. This study aims to test whether the description and processing of a traumatic or highly emotional event in a foreign language could modulate the strength of the connection between traumatic symptomatology and emotional reaction. For this purpose, a sample of 128 healthy participants completed a series of questionnaires via an online platform. Firstly, their levels of distress, arousal and valence were assessed in their native language. Secondly, they were assigned to either the native language or the foreign language group and described a negative childhood event in the assigned language, followed by five questions adapted from Foa and Rothbaum (1998) and related to the event. Next, their emotionality was assessed again in their native language. Finally, a questionnaire of traumatic stress symptoms and an avoidance scale were completed. Multivariate regression analyses showed that the relationship between traumatic symptomatology and emotionality was moderated by the language of processing the negative event. Traumatic symptomatology was more strongly associated with distress and arousal change when the task was performed in the native language. These findings suggest the influence of a foreign language on emotional reactivity when a negative experience is processed, which could be an essential tool in the treatment of disorders related to stress and trauma.

## Introduction

Traumatic symptoms are narrowly associated with an increase in negative emotional intensity (Amstadter & Vernon, 2008), which have been directly related to emotional brain regions such as the amygdala and the hippocampus, as well as areas involving negative intense memories (Jacques et al., 2011). Repressing and storing feelings associated with unpleasant or painful memories has been a key topic in psychological tradition. Unravelling and communicating the distressing information we keep to ourselves has long been considered a healthy habit (Frattaroli, 2006) and is associated with improvements in mental health and recovery (Bistrović et al., 2013). As a method to draw and shape painful past events, emotional disclosure is commonly achieved through a linguistic avenue, either written or verbalized (Pennebaker, 1993; Lepore & Smyth, 2002). However, it is a process that may become resistant due to the patient’s avoidance related to trauma. Also, patients can find the treatment painful, which difficult the dealing with the negative experience. These resistances have resulted in a growing interest in looking for appropriate psychotherapeutic responses in order to improve the experience of treatment (e.g., Chu, 2008, Bicknell-Hentges & Lynch, 2009).

The scientific community has found different ways of helping patients to verbalize their negative experiences, such as drawing (Baker et al., 2017; Hunter, 2019; Malchiodi, 2012) or writing (Sloan et al., 2015). Some authors also have highlighted the use of a foreign language to narrate a negative experience. Different studies have shown that using the native language (L1) is associated with stronger emotional responses compared to a foreign language (L2; e.g., Harris et al., 2003; Pavlenko, 2012). For example, studies in the area of decision-making reflected this difference between languages, showing that the use of one language or the other had an effect on choices and judgments. Keysar, Hayakawa and An (2012) showed how the use of a foreign language diminished the aversion to possible losses in gambles, thus making the decisions more rational and less influenced by emotions. The resulting outcome of this type of experiment gave rise to a pattern that could be interpreted as a reduction in emotionality as well as an increase of rational thinking in a foreign language as compared to the native one, named the ‘foreign language effect’. The more deliberate and rational effect found when using a foreign language in decision-making constitutes a pattern replicated in other tasks within the same field, such as the ones involving moral dilemmas (Costa et al. 2014), taking risks (Hadjichristidis et al., 2015), or inhibiting reluctance to aversive products (Geipel et al., 2018). Other studies also indexed this effect by physiological measures (Eilola & Havelka, 2011; Iacozza et al., 2017).

Although the origin of this effect is unclear, many authors relate the decreased emotionality in L2 either to the context of the acquisition of this language or to the particular processing associated with a foreign language (Caldwell-Harris, 2014). A foreign language is usually acquired in an academic environment in contrast with the early and familiar atmosphere in which the native one was learning. In this ‘colder’ context might grow a larger psychological distance that could evoke less emotionality (Shin & Kim, 2017). Also, Marian and Neisser (2000) specified that the accessibility to the memory was easier when the linguistic context of the recall was the same as the moment of encoding, highlighting the role of language in the process of retrieving autobiographical memories. This is especially relevant framed within the encoding specificity principle in which the properties of a memory can be better retrieved depending on the characteristics surrounding the encoding (Tulving & Thomson, 1973), for instance, the language environment in which it was encoded and retrieved.

A recent study conducted by García-Palacios et al., (2018) showed that conditioning of fear was lighter in a foreign language in comparison to the native one suggesting that the foreign language context can help to reduce emotional reactivity and take distance from the situation. In addition, Ortigosa-Beltrán et al., (in review) explored the effect of a foreign language during the use of cognitive reappraisal and showed that using a foreign language could be advantageous in reducing negative emotionality produced by phobic stimuli by this emotion regulation strategy. In addition, in an experiment on cross-language processing of emotional texts, Dylman and Bjärtå (2018) showed that reading and answering questions regarding negative text extracts in a foreign language was associated with lower distress compared with using the native language. In the same line, Anooshian and Hertel (1994) found a better recall of emotional words in late bilinguals in the native language in comparison to the foreign language, suggesting as a possible interpretation that participants were paying more attention to the pronunciation in the latter. However, other studies did not show this pattern (Ortigosa-Beltrán et al., in press; Ferré et al., 2010).

The clinical setting has intuitively acknowledged this difference between languages from the patients themselves, referencing, in some occasions, a preference for the foreign language. Marcos and his group (Marcos, 1988; Marcos & Alpert, 1976) explored the dynamic of bilinguals in the psychotherapeutic context, finding that Spanish patients verbalizing their experiences in English, their foreign language, expressed a lower level of emotionality. He pointed out that the individuals could be paying more attention to the correct pronunciation and sentence formation in their foreign language than to what they wanted to express, which could be associated with this emotionality reduction. In this regard, Abutalebi (2008) points out that the grammatical and lexico-semantic processing in a foreign language demands more cognitive resources, as shown by the recruitment of cognitive control brain areas, which may be another possible explanation behind this differential effect. Also, Pitta, Marcos and Alpert (1978) emphasized switching languages as an effective treatment for their hysterical clients, which could express their experiences more easily, avoiding the additional emotional charge using their non-dominant language. This effect of using the distance provided by the ‘coldness’ of a foreign language was already noted by Freud and his patients (Freud, 1918) and was later referred to as the detachment effect.

More recently, the foreign language effect has received special attention in the area of recalling traumatic memories, which especially painful to retrieve. Autobiographical memories are composed of sensory information in images, both visual and spatial or auditory, a certain coherence and clarity to a greater or lesser extent, an emotional charge, and language (Greenberg & Rubin, 2003). There is no doubt that language plays a primordial role in the processing of autobiographical memories, making possible the encoding and expression of these memories (Donald, 2012). In this sense, psychotherapy has used language as a tool to rescue negative memories to help deal with those traumatic events that generate discomfort (Larsen et al., 2002). Schrauf, (2000) highlights the language of encoding and retrieval as a marked differential factor and describes a type of pattern in which memories recalled in L2 are remembered more superficially, compared to the high affective content evoked in L1. Studies reported cases of bilingual people who stated that they need to resort to their native language to access the traumatic memories of their childhood more vividly (Aragno & Schlacher, 1996; Javier et al., 1993; Schwanberg, 2010). Iacozza, Costa and Duñabeitia, (2017) also suggest that the use of a foreign language would work as an intermediary or buffer that softens the impact of the emotional distress inherent to strong affective information. In this vein, Jansson and Dylman (2021) also found that using a foreign language modulated the emotionality, vividness and intrusive memories after reactivating emotional autobiographical memories in comparison to a native language. Yet, it is unknown so far to what extent can a different language modulate the relationship between traumatic symptoms related to a negative memory and the emotional response when describing and processing that event.

Therefore, language constitutes a contextual component that can influence the way in which we access and retrieve a memory. The foreign effect in the recall of traumatic memories needs deeper exploration since it may contribute to a better approach in therapy. Patients are commonly reluctant to retrieve emotional events due to the high levels of anxiety and stress when remembering them, leading to the avoidance of thinking or talking about the memory. In this sense, the foreign language effect could function as a tool which could influence how patients process and perceive their emotions. The current study goes in this direction, examining the influence of language in the relationship between posttraumatic symptomatology and emotional reactivity in terms of distress, arousal and valence. We hypothesize that a positive relationship between posttraumatic symptomatology and emotional reactivity will be found, which will be stronger when the negative memory is processed in L1, compared with L2.

## Methods

### Participants

A total of 128 participants (60 women; 63 males; 5 other gender; mean age = 28.70; SD = 9.02) took part in the online questionnaire. All participants were non-clinical and agreed with the informed consent. The inclusion criteria were (1) using Spanish as their mother tongue and (2) having a relatively proficient level of English about their self-perception of knowledge, fluency and use of English (see Table 1), which was measured with a brief questionnaire adapted from the LEAP-Q (Marian, Blumenfeld, & Kaushanskaya, 2007). Differences in terms of English proficiency between participants who performed the task in L1 or L2 were not found (mean native group = 7.84; SD = 1.2; mean foreign group = 7.56; SD = 1.2). The most frequent range of age of foreign language acquisition was between five and ten years old (64%) and also before five years old (17.2%).

**Table 1.** Participant’s basic language skills in the native and in the foreign language g<u>roups (means and standard deviations).</u>

|  | Native (n = 64) | Foreign (n = 64) |
| --- | --- | --- |
| General | 7.84(1.26) | 7.56(1.23) |
| Speaking | 7.20(1.52) | 6.90(1.50) |
| Listening | 8.20(1.22) | 7.58(1.45) |
| Writing | 8.03(1.44) | 7.60(1.40) |
| Reading | 8.73(0.98) | 8.40(1.25) |

### Measures

English skills: **An adaptation** from the LEAP-Q (Marian, Blumenfeld, & Kaushanskaya, 2007) was used to assess self-perception of knowledge of English skills The dimensions assessed were writing, speaking, listening and reading. All items were presented on a scale from 1 to 10. This questionnaire was included in the study in order to control the eligibility and study plausible individual linguistic differences between participants who respond in their native language and those who respond in a foreign language.

Post traumatic symptomatology: The Davidson Trauma Scale (DTS; Davidson et al., 1997) is a 17-item questionnaire to self-report the frequency and severity of PTSD symptoms, ranging between “not at all” and “every day” on a 5 point scale. The questions refer to the symptoms of the chosen negative event in the past week. However, participants were asked to answer the questions of this scale regarding their negative childhood experiences recalled in the previous part of the experiment.

Distress: The subjective Units of Distress Scale (SUDS; Wolpe, 1969) was used following Dylman and Bjärtå (2018) as a measure of distress and discomfort, ranging from 0 (no distress) to 10 (high distress). Previous experiments used it as a scale of distress and anxiety (Tanner, 2012). This scale also correlates with the State Trait Anxiety Inventory (STAI; Spielberger, Gorsuch, Lushene, Vagg & Jacobs, (1983).

Valence and Arousal: The Self-Assessment Manikin (SAM; Lang, 1980) was administered to assess ratings of valence of their emotions and the self-report of arousal (people’s reports of affective experience). The scale of valence ranged between 1 (positive valence) and 5 (negative valence), and the scale of arousal ranged also between 1 (high arousal) and 5 (low arousal).

### Design and procedure

This is a cross-sectional study in which participants completed an online survey displayed with the online platform Qualtrics (Qualtrics, L. L. C., 2010) and disseminated by Prolific (www.prolific.co). The study was approved by Universitat Jaume I ethical committee (CD/100/2021). Participants entered to the platform according to the criteria related to the native language (Spanish), country of birth and residence (Spain) and a general estimate of the level of English (good/high level). When the eligibility was confirmed, they were informed of the consent form and asked to provide their email, age, and gender. The emails prior experiment and the information on how to complete the tasks were in the native language in order to discard a possible influence of L2. Then, participants completed the English questionnaire, adapted from the LEAP-Q (Marian, Blumenfeld, & Kaushanskaya, 2007), to measure the specific self-perceived level in each of the English dimensions (speaking, listening, writing and reading). Next, they reported the previous levels of distress and emotional affection in order to establish baseline levels of distress, valence and arousal. After that, half of the participants proceeded in their native language (Spanish) and the other half in their foreign language (English). They were requested to think briefly and describe a negative event of the past in the corresponding language, following the memory recall paradigm of free recall used previously (Otoya, 1987; Slofstra et al., 2017). Specifically, they were asked to describe a painful, negative or traumatic event of their childhood following previous literature so that all of them referred to the same period of their lives. Then, the participants had to rate the emotionality of the event on a scale from 0 to 10 and the level of interference at the present time. Next, the five processing questions related to the event were shown in the corresponding language. The processing questions were adapted from the protocol by Foa and Rothbaum (1998) for post-traumatic stress disorder (PTSD). The description of the event works as part of the processing here, taking into account that the recall of an autobiographical memory could increase the negative affect (Slofstra et al. 2017). In the last phase, all participants returned to the native language and completed the same questionnaires as the first phase to determine the level of distress, valence and arousal. They also completed the DTS at the end of the task in order to not affect the emotional intensity during the processing task. The payment was made through the platform and adjusted to the time spent by each participant (9 euros per hour).

### Data analysis

To check the effectivity of the processing task increasing emotionality, three t-student tests were performed with pre and post-scores of all the dependent variables (distress, valence and arousal). Once changes in emotionality were checked, differences between these pre and post-processing scores were calculated to obtain different measures of change in emotionality. Differences were calculated by subtracting the value pre-processing from the post-processing value. Thus, higher change values on SUDS and valence mean more emotional intensity in terms of distress and displeasure, while lower change values on arousal indicate more emotional intensity. Next, bivariate correlations were analyzed in order to study the association between the study variables (post-traumatic stress symptomatology, distress change, arousal change, valence change and Language). To test the moderating effect of Language in the relationship between post-traumatic stress and emotionality, multiple linear regressions were used via the macro PROCESS (Hayes, 2017). Specifically, 3 regressions were performed, one with each dependent variable (distress, arousal and valence), post-traumatic stress symptomatology as the independent variable, and language as the moderator. Conditional effects of the independent variables on the dependent variables were obtained, as well as a graphical representation to interpret the findings. An alpha level of 0.01 was set for all analyses to reduce Type I errors.

## Results

### Emotional reactivity during the processing task

T-test showed that the processing task was effective inducing emotional reactivity, showing higher arousal (t=4.80; p<.0001), higher distress (t=4.89; p<.0001) and more displeasure (t=8.15, p<.0001) during the post-processing compared with the pre-processing phase.

### Bivariate correlations between study variables

Pearson bivariate correlations showed that post-traumatic stress symptoms were positively associated with changes in both distress and valence (distress: r=.27, p=.009; arousal: r=.24, p=.009), while was negatively associated with change on arousal (r=.23, p=.012). Language was not associated with any of the other study variables (Table 2).

**Table 2.**
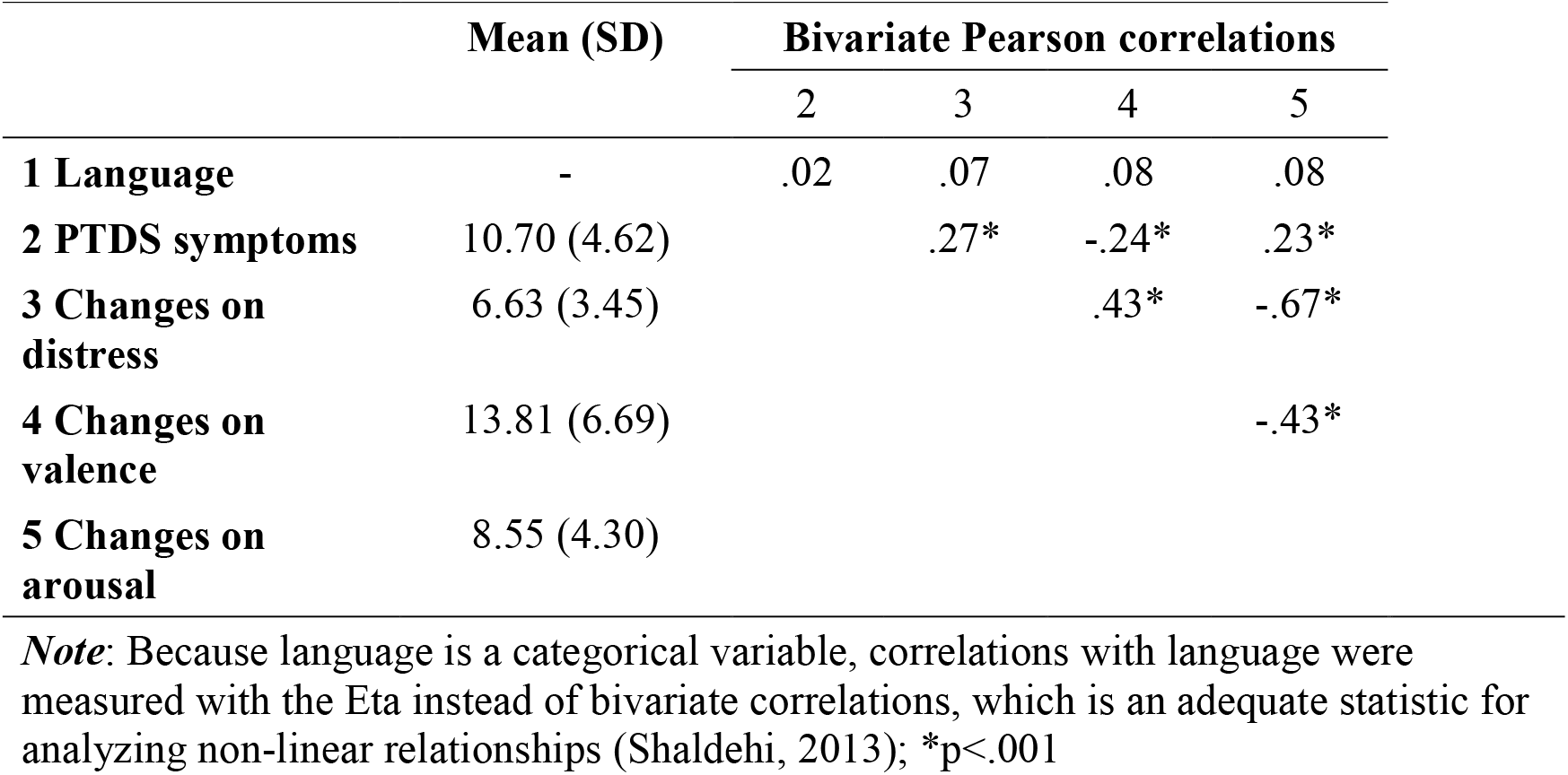
Bivariate correlations between language, post-traumatic stress disorders, and <u>changes on distress, valence, and arousal</u>

|  | Mean (SD) | Bivariate Pearson correlations |  |  |  |
| --- | --- | --- | --- | --- | --- |
|  |  | 2 | 3 | 4 | 5 |
| **1 Language** | - | .02 | .07 | .08 | .08 |
| **2 PTDS symptoms** | 10.70 (4.62) |  | .27* | -.24* | .23* |
| **3 Changes on distress** | 6.63 (3.45) |  |  | .43* | -.67* |
| **4 Changes on valence** | 13.81 (6.69) |  |  |  | -.43* |
| **5 Changes on arousal** | 8.55 (4.30) |  |  |  |  |

*Note:* Because language is a categorical variable, correlations with language were measured with the Eta instead of bivariate correlations, which is an adequate statistic for analyzing non-linear relationships (Shaldehi, 2013); \* $p < .001$

### Moderation of language

Results from the multivariate regression analysis are shown in the Table 3. The main effect of post-traumatic stress symptoms was significant for distress (β=0.02, t=-3.92 p=.0002, 99% CI = [0.01, 0.03]).), arousal (β=-0.01, t=-3.29, p=.0013, 99% CI = [0.63, 0.01]) and valence (β=-0.01, t=2.57, p=.0115, 99% CI = [0.002, 0.018]). Language main effect was not significant for any model. Regarding the moderation effects, results showed that the interaction post-traumatic stress symptomatology x language was significant in fostering distress (β=-0.02, t=-2.45, p=.016, 99% CI = [-0.038, -0.040]) and arousal (β=0.01, t=2.02, p=.045, 99% CI = [0.002, 0.023]), while was not significant for valence. As shown in Table 4, the analyses of conditional effects showed that the strength of the relationship between post-traumatic stress symptomatology and emotional reactivity (distress and arousal) was stronger in participants who do the emotional processing in their native language, while this relationship decreased, being nonsignificant when participants recall and process this memory in a foreign language. A graphical representation of the relationship between post-traumatic stress symptomatology and distress and arousal depending on the processing language is shown in Figure 1 and Figure 2, respectively.

**Table 3.** Multivariate hierarchical regression analyses predicting distress, valence and arousal changes from post-traumatic stress symptoms, language, and their interaction

|  | Distress |  |  |  |  | Valence |  |  |  |  | Arousal |  |  |  |  |
| --- | --- | --- | --- | --- | --- | --- | --- | --- | --- | --- | --- | --- | --- | --- | --- |
| | $\beta$ | t | p | 99% CI | R<sup>2</sup> | $\beta$ | t | p | 99% CI | R<sup>2</sup> | $\beta$ | t | p | 99% CI | R<sup>2</sup> |
| PTSD symptoms | .02 | 4.92 | .0002 | .011, .033 | .07 | .01 | 2.57 | .012 | .002, .018 | .13 | -.01 | -3.30 | .001 | -.020, -.005 | .10 |
| Language | -.14 | -0.68 | .501 | -.565, .278 |  | -.14 | -.95 | .342 | -.434, .162 |  | .13 | .95 | .346 | -.148, .419 |  |
| PTSD symptoms x<br>Language | -.02 | -2.45 | .016 | -.038, -.004 |  | -.005 | -.83 | .409 | -.017, .007 |  | .01 | 2.02 | .046 | .0002, .023 |  |

**Table 4.** Conditional effects language in the relationship between post-traumatic stress symptoms and distress and arousal

| **Catastrophizing component-**<br>**Dependent variable combination** | **Language** | **$\beta$** | **$t$** | **$p$** | **99% CI** |
| --- | --- | --- | --- | --- | --- |
| PTSD symptoms-distress | Native | .02 | 3.922 | .0002 | .011, 0.034 |
|  | Foreign | .001 | 0.178 | .859 | -.012, 0.014 |
| PTSD symptoms-arousal | Native | -.01 | -3.299 | .001 | -0.020, -.005 |
|  | Foreign | .001 | -.197 | .844 | -0.010, .008 |

**Figure 1.**
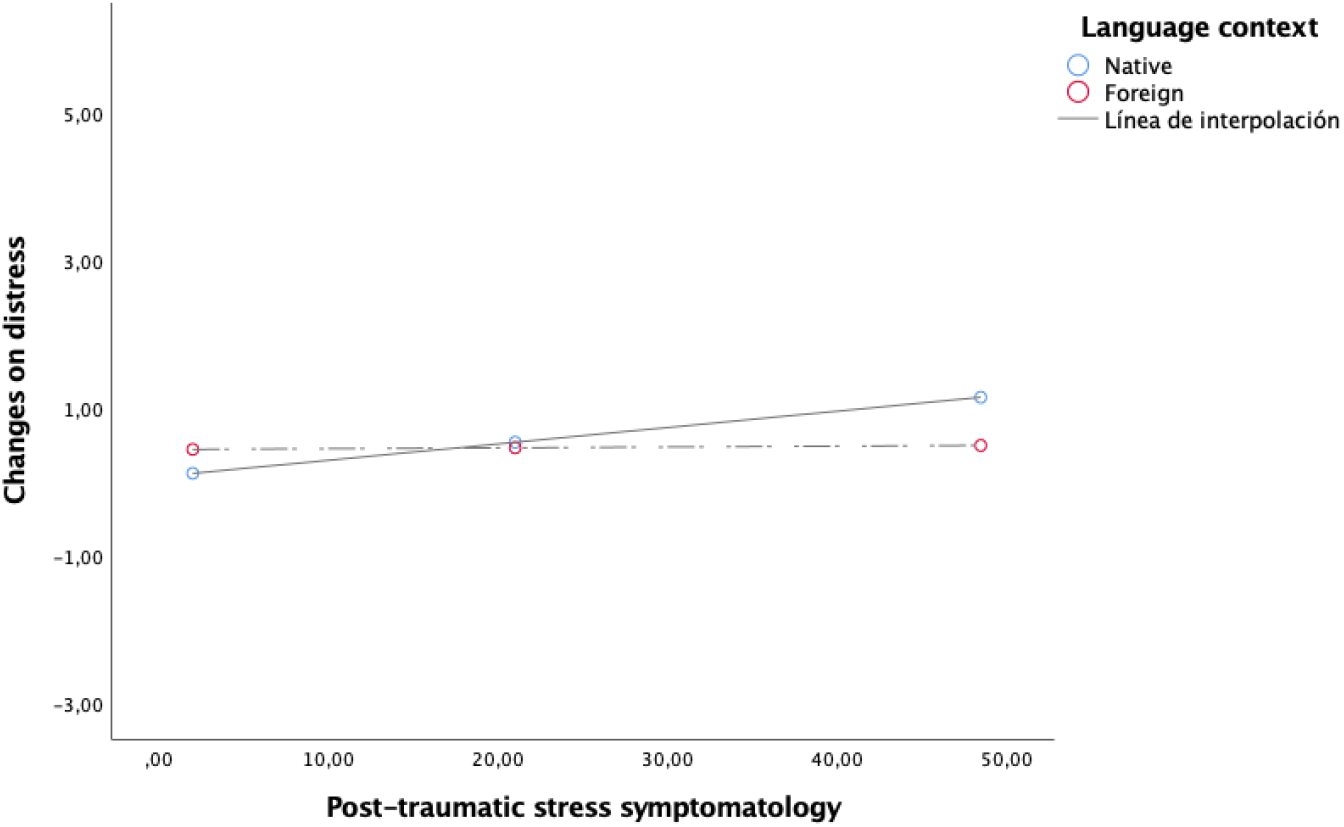
Score changes between these pre and post on distress.

**Figure 2.**
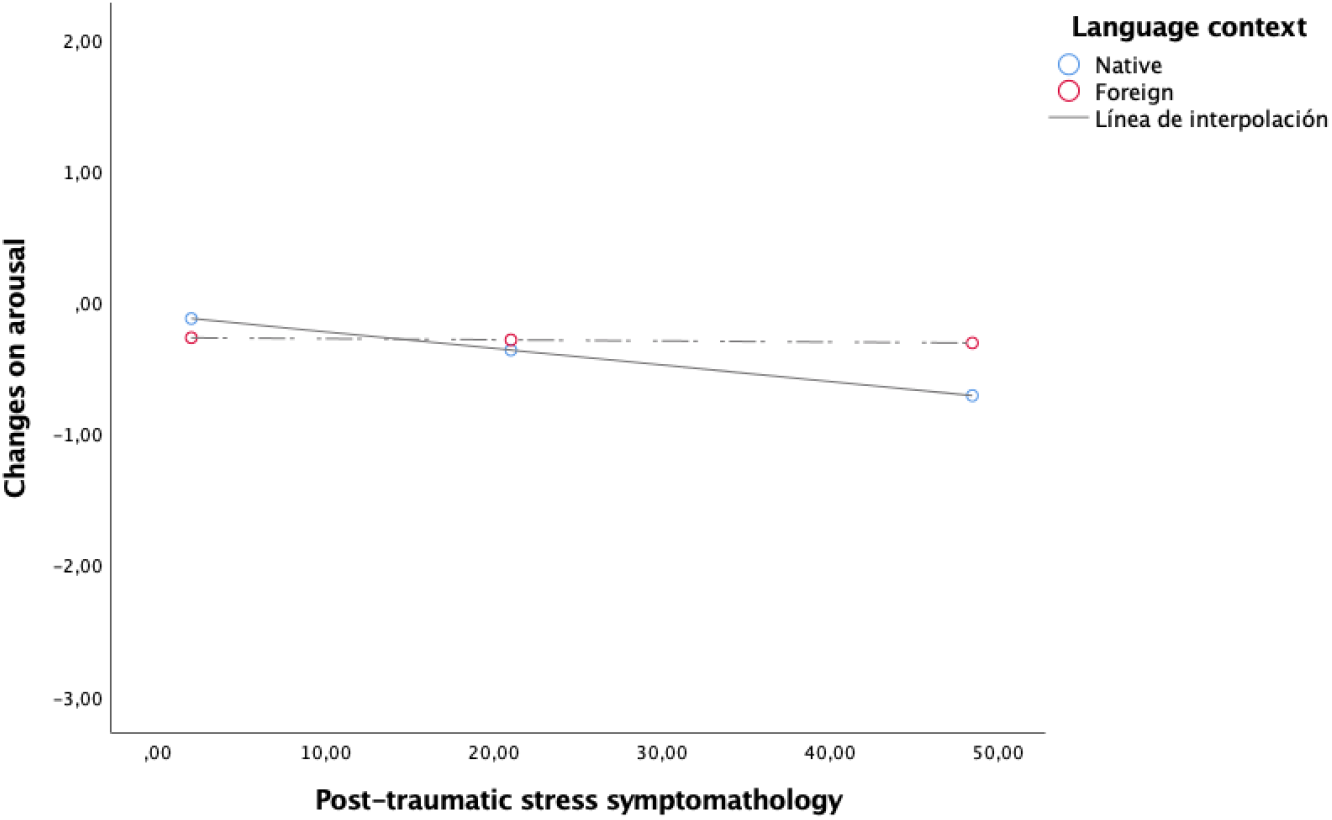
Score changes between these pre and post on arousal.

## Discussion

The present study is the first to explore whether the language of retrieval modulates the connection between trauma symptoms and the level of emotionality, in particular, distress, valence and arousal, when processing a negative autobiographical event.

As proposed in the initial hypothesis, the results showed that the relationship between the strength of traumatic symptoms and self-reported emotionality, specifically distress and arousal, is moderated by the language used. In particular, this strength is greater when the native language is used compared with using a foreign language. These results are consistent with previous studies that support that the use of a native language produces a stronger emotional response than using a foreign language (Pavlenko, 2012; Iacozza, et al., 2017). These results are explained in terms of attentional resources, which could be divided between using a language that is not mastered and the remembered event, resulting in less focus on the event itself and consequently working as a psychological or emotional barrier when managing the emotionality that the processing provokes. Thus, in line with Anooshian and Hertel (1994), we highlight the importance of attention in the modulation of the emotional processing of negative events.

These results have important clinical implications for bilingual patients. As previously mentioned, some patients are reluctant to address their negative memories due to the high levels of anxiety and stress when remembering them. On the base of it, it seems feasible here to assume support for the case studies that posited a native language as a facilitator to access past negative events, in contrast to the foreign language (Marian & Neisser, 2000). This access to a memory in a foreign language would undermine the connection between traumatic symptoms and the resulting emotional reaction. Hence, language may be an interesting tool when psychotherapists need to decrease emotionality to do a first approximation to the negative experience.

However, the use of a foreign language could also have some inconveniences in psychotherapy. In view of our findings, it is important that psychotherapists take into account the role of language as a moderator in the relationship between trauma symptomatology and emotional reactivity when working with bilingual or multilingual patients. The use of a foreign language could entail a higher detachment during the processing of a negative event, which could difficult some psychotherapeutic processes such as debriefing. In fact, literature commonly signaled more structured past memories in L1 with richer descriptions (Javier & Marcos, 1989), as well as more memories when cued with words in the native language (Otoya, 1987).

This study has some strengths since it opens the door to a different approach to work with negative autobiographical memories, in which language plays a primary role during the psychotherapeutic process. These findings support the idea by Dewaele and Costa, (2013) of the need to pay more attention to language as an influent factor within the multilingual clinical context. Patients can use bilingualism or multilingualism as a tool to adjust their needs of communication in therapy, as discussed by Dewaele et al., (2020). Even though we have not explained the phenomenon of the foreign language effect, we can bring some perspective to how it works in relation to negative memories and their processing, and ultimately the modulatory role this effect has on emotional response. Still, different paradigms and tasks remain to be explored, especially in relation to negative charged events. There are some considerations to have into account in this study. One may be the recruitment of a larger sample. As this experiment explores healthy population, the scores are concentrated in low scores in traumatic symptomatology, and the differences are clearer when the DTS scores are higher, so it is recommended to conduct a study with a clinical population either with trauma or with people with an adaptive disorder who have gone through a stressful event. In relation to language, it would be appropriate to specify weather language differences are due to the language itself or to language switching, as in this particular task participants in the foreign language group had to switch from their native language to their foreign for a few items. Recent research has already pointed out the fact that switching from the native language to the foreign might lead to a facilitation in the processing of negative responses (Zang et al., 2022). In addition, although participants were asked to recall memories from childhood, the majority of the memories were experienced in adulthood; this could imply different outcomes depending on the distance between the memory and the period in which it was experienced. Recent memories, for example, are typically perceived as more vivid and emotionally intense compared to remote memories (Sutin & Robins, 2007).

All in all, we cannot confirm with these results that a different language directly evokes a different emotional intensity. However, these findings have shown that retrieving a negative past event in a foreign language modulates differently the emotional reaction associated with traumatic symptomatology. With this, we tentatively suggest that a foreign language can modulate the emotional expression of certain aspects of PTSD symptoms.

In conclusion, our results are consistent with prior results of bilingual memory, as well as with those exploring emotional material in a foreign language. These findings shed light on the modulatory function that language has with respect to the retrieval of emotional events of the past and raise new questions on the field of recalling negative memories.

## Data Availability

All data produced are available online at Open Science Framework (OSF)

https://osf.io/n4wvk/

## Author contributions

AG-P, AC and IO-B developed the concept of the study and provided ideas for the study design alongside with IJ. IO-B prepared the questionnaires in the online platform. IO implemented the data collection and the testing of the participants. IJ executed the data analysis and interpretation of the results alongside with IO-B. IO-B elaborated the manuscript with IJ under the supervision of AG-P.

## Acknowledgments

The authors would like to thank the Multilingual Mind project for its support in every way and for providing all the facilities.

## Availability of materials

The datasets analyzed for this study can be found in the Open Science Framework (OSF) Repository [https://osf.io/n4wvk/].

